# Analytical solution of equivalent SEIR and agent-based model of COVID-19; showing the bounds of contact tracing

**DOI:** 10.1101/2020.10.20.20212522

**Authors:** Huseyin Tunc, Fatma Zehra Sari, Busra Nur Darendeli, Ramin Nashebi, Murat Sari, Seyfullah Kotil

## Abstract

Mathematical models not only forecast the possible future but also is used to find hidden parameters of the COVID-19 pandemic. Numerical estimates can inform us of both goals. Still, the interdependencies of parameters stay obscure. Many numerical solutions have been proposed so far; however, the analytical relationship between the outbreak growth, decay and equilibrium are much less studied. In this study, we have employed both an equivalent agent-based model and a Susceptible-Exposed-Infected-Recovered (SEIR)-like model to prove that the growth rate can be determined analytically in terms of other model parameters, including contact tracing rate. We identify the most sensitive parameters as undocumented transmission rate and documentation ratio. Unfortunately, these are the parameters we have the least knowledge. We derived an identity that predicts the effectiveness of contact tracing in a country from observable parameters. We underline an unavoidable dilemma: that even in the case of high contact tracing, we cannot bring the outbreak to stalemate without applying substantial quarantine; however, some countries are benefiting from contact tracing. Besides, we have shown that the seemingly same parameters of the SEIR models and agent-based models are not equivalent. We propose a correction to bridge both models.

## Introduction

In December 2019, a novel enveloped RNA beta-coronavirus that causes coronavirus disease (COVID-19) emerged in Wuhan, China, and the disease has become a global crisis rapidly. Besides being a global health-threatening factor, recent studies have shown that economic spillovers and spillbacks will be very large^1^, each additional month of the epidemics costs 2.5-3% of global GDP^2^, an increment of global poverty will be expected^3^. Therefore, the issue of finding efficient strategies to prevent the spread of the disease becomes more and more prominent for policymakers.

Agent(individual)-based models and equation-based models are two common frameworks to investigate the dynamics of epidemics and the efficiency of the prevention strategies. The way of modelling the relationships between entities and the level-of-detail are the points where these two approaches differ. Agent-based models can capture individual contact processes and give a more realistic view of an outbreak and its evolution, however, the advantages come with costs of heavy computational burden and time to run the simulation. In standard compartmental models that are governed by a system of differential equations, the population is split into homogenous subpopulations such as S-[E]-I-R (Susceptible-[Exposed]-Infectious-Recovered/Removed). Despite their short set up and running time, the homogeneity assumption is a shortcoming of compartmental models due to the small number of infective individuals at the beginning of the disease and the stochastic nature of transmission^4^. Branching processes are good approximations to the stochastic epidemic processes at the onset of epidemics when there are few infected individuals, and the number of susceptibles is large^5^.

Even though agent-based modelling for COVID-19 is yet relatively rare, studies such as taking different transmission rates of symptomatic and undocumented patients into account and estimating population compliance level and making comparisons of different intervention strategies^6^, estimating the effectiveness of measures via taking super-spreaders and testing and quarantine-policies into account^7^, constructing a virtual community and calculating the effect of delay-time for interventions^8^ were published. There are also studies that use the agent-based modelling for combining human mobility data with SIR-like model and calculating the effect of measures on the peak level^9^, using micro agent-based modelling and estimating the effect of interventions on the disease’s cumulative incidence and mortality, and on ICU-bed occupancy^10^, considering different stages of the disease and estimating the number of patients in hospitals and ICU^11^, dividing society into groups and estimating cost vs quarantine length^12^.

Since the beginning of the epidemic, many statistical, mathematical, and agent-based models have been used to estimate the epidemiological parameters and effectiveness of intervention strategies. However, limited knowledge about the nature of the disease, methodological differences(choice of parameters), model differences(deterministic versus stochastic) and use of different datasets caused asymptomatic cases were missed^13-14^, wrong predictions about the expected number of infected individuals^15-16^ or negligence of time delay between symptom onset - infectious state^17-19^.

Previous works investigated the effects of intervention strategies, including contact tracing^20-26^, quarantine^8,9,21,24-29^, and testing^30,25-26^ on epidemiological parameters of the disease. As a result of investigating these interventions separately, the question to be answered to implement the most effective quarantine intervention; when and how long the quarantine is applied, the question to be answered for contact tracing; how many people contact tracing can be done on, for the random testing; determining whether the test application has an impact on the number of cases and the cost of the measures. When applied separately, it was observed that they have difficulties in practice^20^, although the effect of contact tracing and quarantine was demonstrated in reducing epidemic spread.

Here, we have analytically derived a new equation which shows the relation of internal parameters of COVID-19 pandemic and the external control parameters. The derivation was made from an SEIR-like model that we constructed to be equivalent to our agent-based model. The models also include contact-tracing. We show that for both models to be equivalent, we must make corrections on the coefficients. The constructed models and our solution are modelling the exponential phase of the epidemic. The populations with low herd-immunity are in the exponential phase of the dynamics. Thus, practically all countries are in the exponential growth or decay phase. We then identified the most sensitive and correlating parameters on the pandemic growth rate. Through our solution, it is possible to find out the ratio of the intervention strategies to reach the equilibrium state. We show that increasing effectiveness of contact tracing does not have a substantial effect on the growth rate. We have also created an analytical expression which can predict the efficiency of contact-tracing from observable parameters. Also, we can compute the proportion of new cases originating from contacts that were identified earlier (or in isolation). Lastly, we have proved that the uncorrected SEIR-like models lead to the miscalculation of day estimates. We have confirmed all of our findings with the agent-based models.

## Results and Discussion

### Mathematical modelling

We have developed a discrete-time stochastic agent-based model and corrected SEIR-like model, including contact tracing strategies parameterized to the COVID-19 outbreak. The dynamic states include *E*(*t*) (exposed), *P*(*t*) (pre-symptomatic documented), *U*(*t*) (undocumented), *D*(*t*) (symptomatic documented), *H*(*t*) (hospitalized) and *R*(*t*) (recovered) individuals as shown in Fig. 1. By assuming exponential growth/decay of all state variables, we have proved that the people who will leave his/her own state in the classical SEIR model Eq. (15) cannot be distributed uniformly (see methods). With the help of our mathematical analysis over the agent-based model, we have proved that the coefficients like 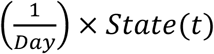 occurred in classical SEIR model must be replaced with the corrected coefficients 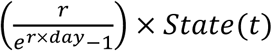 where *r* is corresponding growth rate. Thus, we have derived a corrected SEIR-like model Eq. (18) which is equivalent to the mean behaviour of the agent-based model. The amount of error that will occur in the day predictions made with the uncorrected SEIR models an be given in the following form,

**Figure 1.**
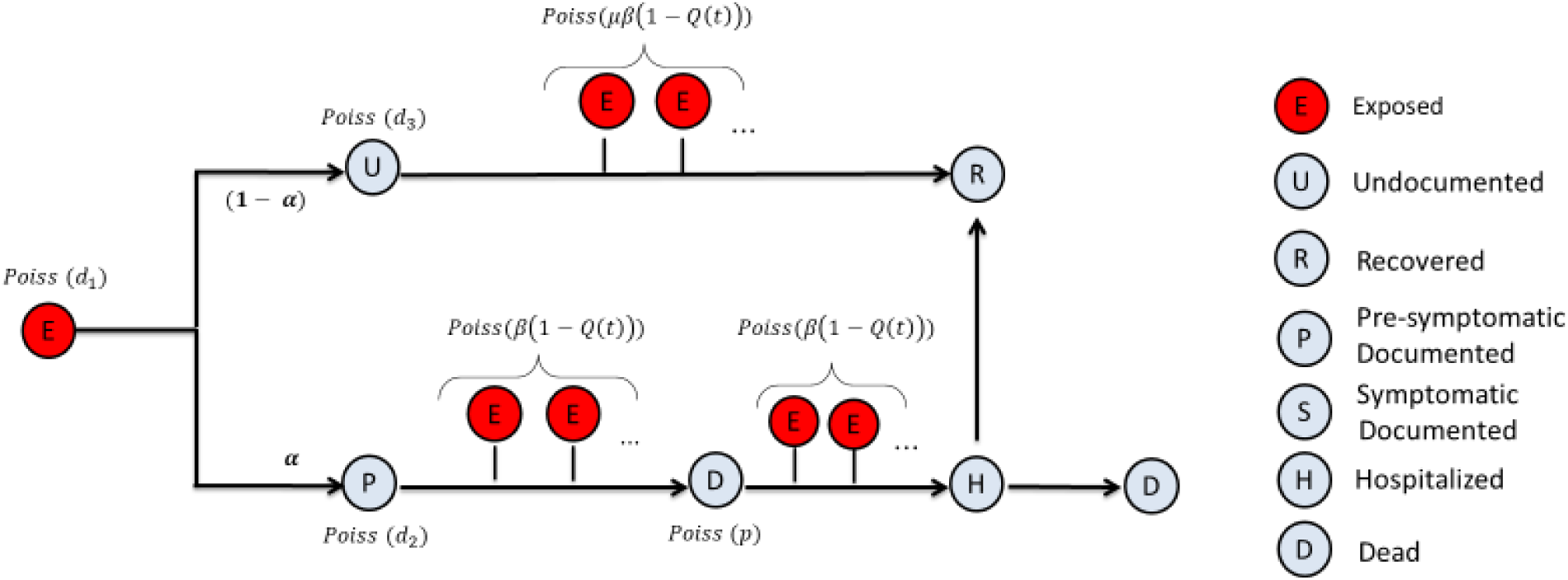
Graphical representation of the models and the parameters. Every COVID-19 positive agents start in the exposed state. Then, exposed individuals proceed randomly to undocumented or pre-symptomatic documented states. Different transmission rates of pre-symptomatic and undocumented cases have been taken into account. The susceptible agents are omitted due to low-herd immunity limit. The new exposed individuals are added as Poisson random variables per infectious patients. A single parameter, Q, has been used to represent all precautions implemented to prevent the spread of the disease.

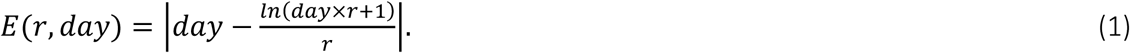

One major challenge in SEIR-like models is the addition of contact tracing strategies effectively. The corrected SEIR-like model is extended to include contact tracing strategies by considering isolated compartments as well. The derived model is given by Eq. (20). As we will prove through the simulations, the corrected SEIR-like model that includes contact tracing is also equivalent to the agent-based model for which the contact tracing strategies is explained in the methods. With the mathematical analysis of our corrected SEIR-like model Eq. (20), we have derived the following generalized identity

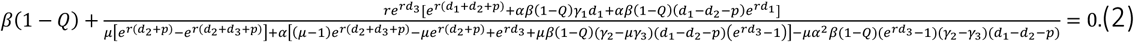

where the default parameter values and their roles are given in Table 1. The equilibrium identity can then be derived from Eq. (2) as follows,

**Table 1.** Parameters used in the model. Symbols, descriptions and values of the model paramaters.

| Parameter | Definitions | Values | Source |
| --- | --- | --- | --- |
| $\alpha$ | the fraction of documented infections, | 0.14 | [31] |
| $d_1$ | mean number of days in exposed stage | 3.69 | [31] |
| $d_2$ | mean number of days in pre-symptomatic documented stage | 3.47 | [31] |
| $d_3$ | mean number of days in the undocumented stage | 3.47 | [31] |
| $p$ | average time of going to the hospital | 1.92 | [34] |
| $\beta$ | infectiousness rate of the symptomatic case | 1.12 | [31] |
| $\mu$ | transmission reduction factor for undocumented cases | 0.55 | [31] |
| $Q(t)$ | level of total precautions in a country | Depends on sim. | Estimated |
| $\gamma_1$ | isolation probability of contacts at the exposed stage | Depends on sim. | Assumed |
| $\gamma_2$ | isolation probability of contacts at the pre-symptomatic documented stage | Depends on sim. | Assumed |
| $\gamma_3$ | isolation probability of contacts at the undocumented stage | Depends on sim. | Assumed |

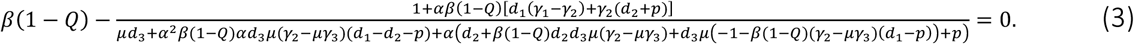

One more identity is derived for calculation of known-ratio, which is the proportion of new daily identified patients as traced from earlier contacts. The known-ratio is obtained as

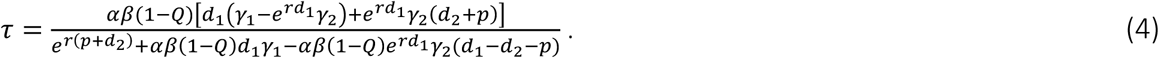

### Accuracy of the analytical solution compared to the agent-based model

We test the accuracy of the novel Eq. (2) with our agent-based model in terms of the logarithmic growth rates in Fig. 2. The comparison has been performed for all parameters, including contact tracing, and it has been shown that the novel equation predicts the growth rate with high accuracy. Even though the stochasticity of agent-based modelling has caused minor deviations between the novel equation and the agent-based model, the effect of the parameters on the growth rate has been found similar with only a 1% difference between the two models. Therefore, the proposed novel equation can be used to analyze the dynamics of the epidemics with respect to model parameters.

**Figure 2.**
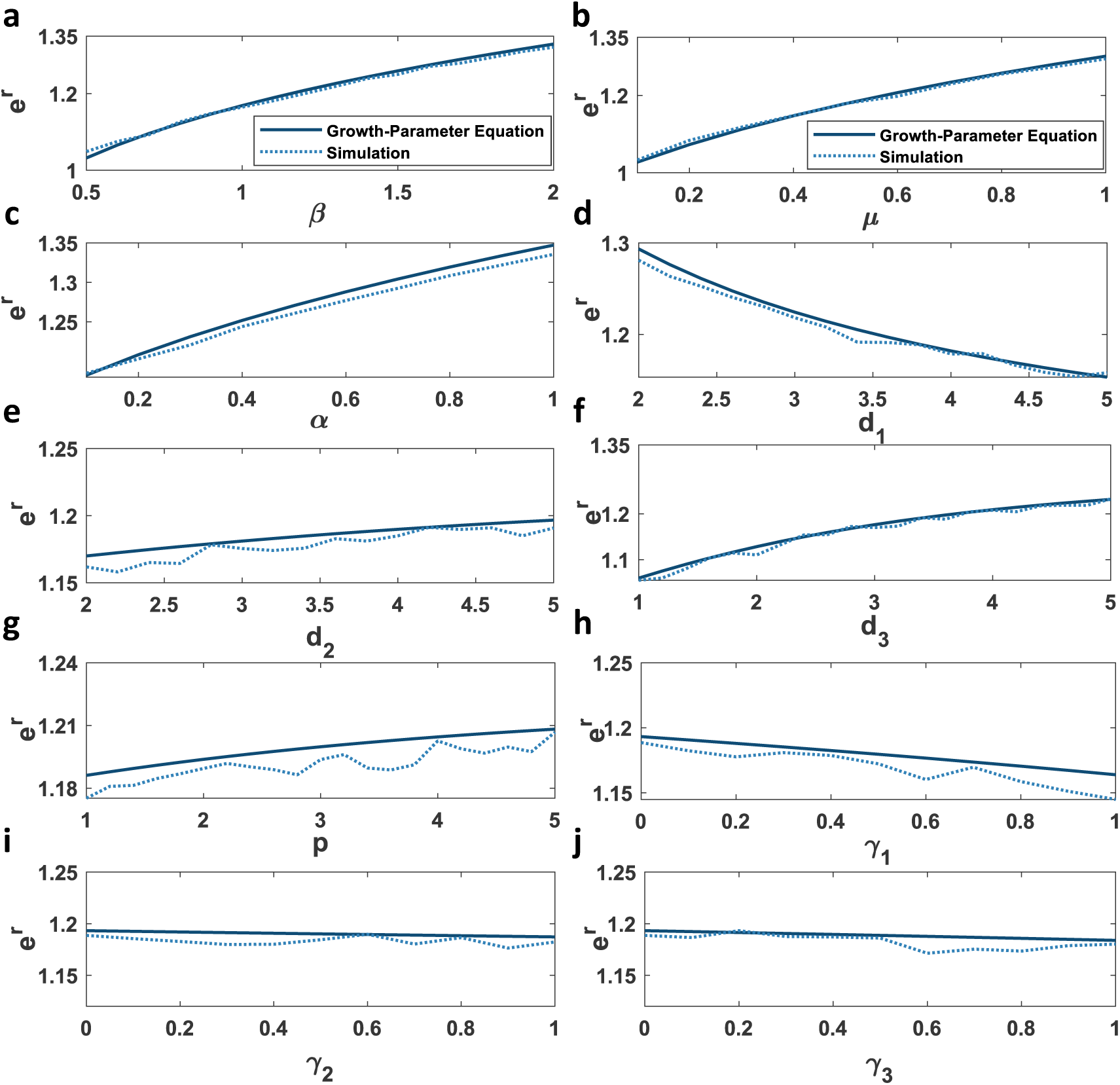
Analytical solution predicts the growth rate of the agent-based model. The comparison of the novel equation and agent-based modelling has been performed with respect to **a**, infectiousness rate of the symptomatic documented cases *β* **b**, transmission reduction factor for undocumented cases *μ* **c**, the fraction of documented cases *α* **d**, the mean number of days in exposed stage *d*_1_ **e**, the mean number of days in the undocumented stage *d*_2_ **f**, the mean number of days in the pre-symptomatic documented stage *d*_3_ **g**, the mean number of days in the symptomatic documented stage *p* **h**, isolation probability of contacts at the exposed stage *γ*_1_ **i**, isolation probability of contacts at the pre-symptomatic documented stage *γ*_2_ **j**, isolation probability of contacts at the undocumented stage *γ*_3_. (see methods parameter values)

### Correlation and sensitivity analysis of the COVID-19

The linear correlations of all parameters with the growth rate of the epidemic are illustrated in (Fig. 3 a). The transmission rates of both undocumented and symptomatic cases (*β, μ, α*) have shown a dominant positive correlation with the growth rate of the disease. Another dominant parameter is the latency period. The length of the latency period is inversely correlated to the growth rate. In contrast, the length of the infectious period is positively correlated. The analysis also includes contact tracing. We divided the efficiency of contact racing into three parts (Table 1). Isolation of contacts at the latency period is more effective than to isolate them when they are at pre-symptomatic or undocumented stages to reduce the growth rate of the disease. The contacts that do not show any signs of disease are the hotbed of the spread of the epidemic. They must certainly be quarantined/isolated.

**Figure 3.**
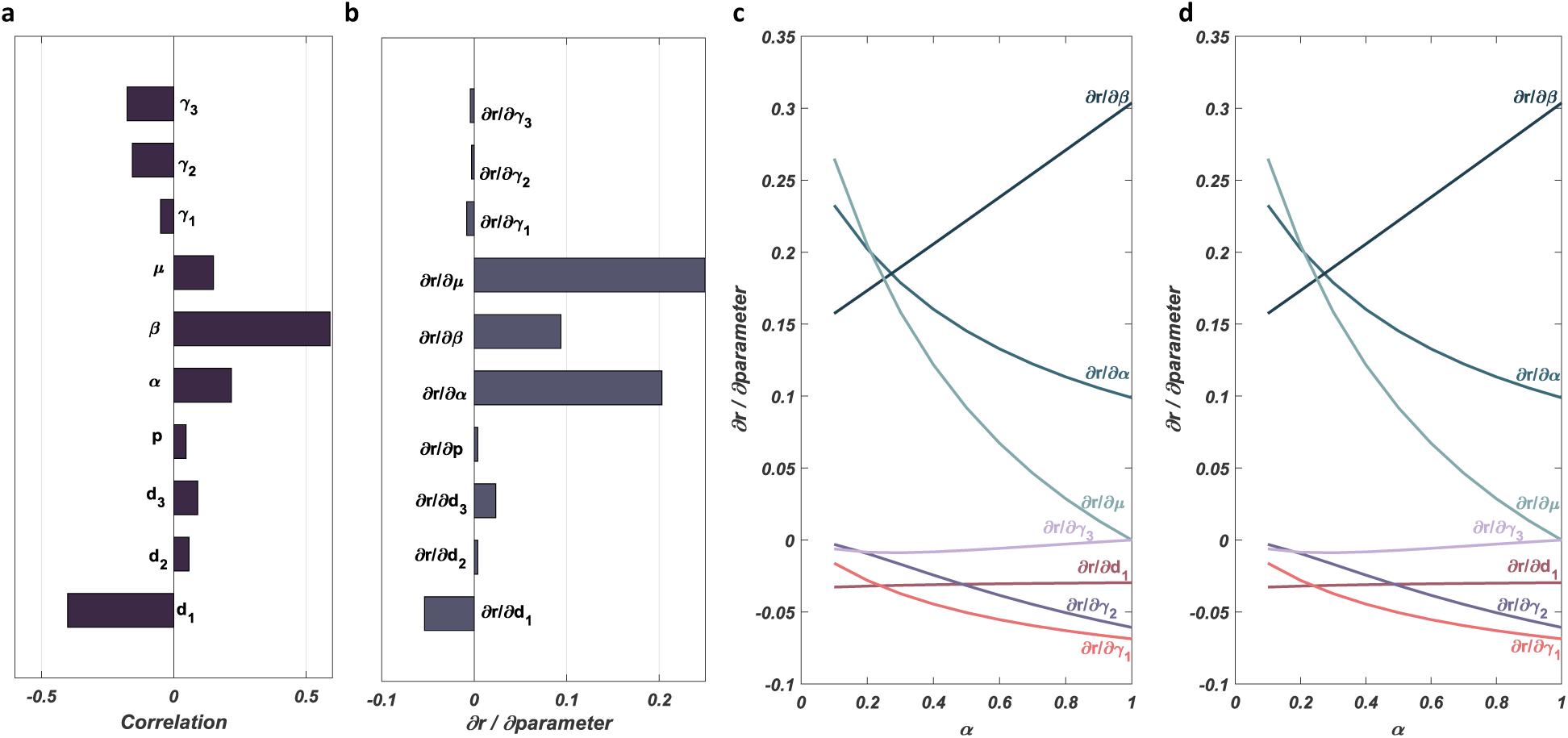
Correlation and sensitivity analysis of the COVID-19 dynamics shows the importance of non-observable parameters. Correlation of the parameters with growth rate and sensitivity analysis by considering default parameters stated in Table 1. **a**, Correlation between the model parameters and the growth rate of the pandemic has been determined by using the novel Eq. 2 **b**, Sensitivity analysis of the growth rate of the pandemic calculated via partial derivatives of the growth rate with respect to the parameters **c**, Sensitivity analysis with the use of default parameters and contact tracing probabilities *γ*_1_ = *γ*_2_ = *γ*_3_ = 0 for various values of the documentation ratio *α*. **d**, Sensitivity analysis with the use of default parameters and contact tracing probabilities *γ*_1_ = *γ*_2_ = *γ*_3_ = 1 for various values of the documentation ratio *α*. (For the derivation, Eq. (2) and default parameters are used)

Sensitivity analysis has been performed to find out the disease’s growth rate is more sensitive to which parameter of the model (Fig. 3 b). The sensitivity is not synonymous with correlation. The sensitivity shows us the effect of the uncertainty of the parameters. Especially four parameters have been found as more sensitive among all parameters. Those parameters are the transmission rate of documented cases (β), documentation ratio (α), the transmission reduction factor of the undocumented cases (*μ*), and the mean number of days in the latency period(*d*_1_). The most sensitive parameter is the transmission reduction factor of the undocumented cases (*μ*), which is one of the most controversial parameters about COVID-19^33^. Depending on the various values of the documentation ratio *α*, sensitivity analysis is performed to illustrate the different scenarios in (Fig. 3 c and 3 d). The sensitivity of the growth rate with both tracing-free case and the full-tracing case has been analyzed in Fig. 3 a, and Fig. 3 b, respectively.

As we observe from Fig. 3 c, the sensitivity of the growth rate to the transmission rate (*β*) is increasing with increasing values of the documentation ratio. It indicates that the transmission rate is the dominant parameter when the documentation ratio is high as opposed to our default case illustrated in (Fig. 3 b). For higher values of the documentation ratio, the sensitivity of the growth rate to the transmission reduction factor of undocumented cases (*μ*) is getting insignificant. For high contact tracing, it has been observed that the documentation ratio is not a dominant parameter (Fig. 3 d) as opposed to our default case stated in (Fig. 3 b). An interesting point here is that when the tracing values and documentation ratio are relatively high, a small positive change in documentation ratio leads to an adverse change in growth rate. The reason for this change is the increase in documentation ratio increases the effect of tracing. On the other hand, tracing parameters (*γ*_1_) and (*γ*_2_) become dominant parameters in terms of the sensitivity in case of higher values of the documentation ratio and higher values of contact tracing probabilities (Fig 3 d).

### Quarantine/Precaution level required to stabilize the disease spread

We estimate the threshold quarantine level that can equilibrate the pandemic in the presence or absence of contact tracing, *Q*_*E*_ and *Q*_*T*_ respectively (Fig. 4 a and 4 b). The quarantine level is the sum of all precautions like masks, social distancing, isolation. Any quarantine level higher than the threshold will turn the epidemic into decay. Afterwards, we compute the quarantine level for exponential decay with decay rate r = −0.2 (Fig. S4 a and S4 b). Contact tracing cannot make a significant effect to decrease the threshold quarantine levels at the low documentation ratio.

**Figure 4.**
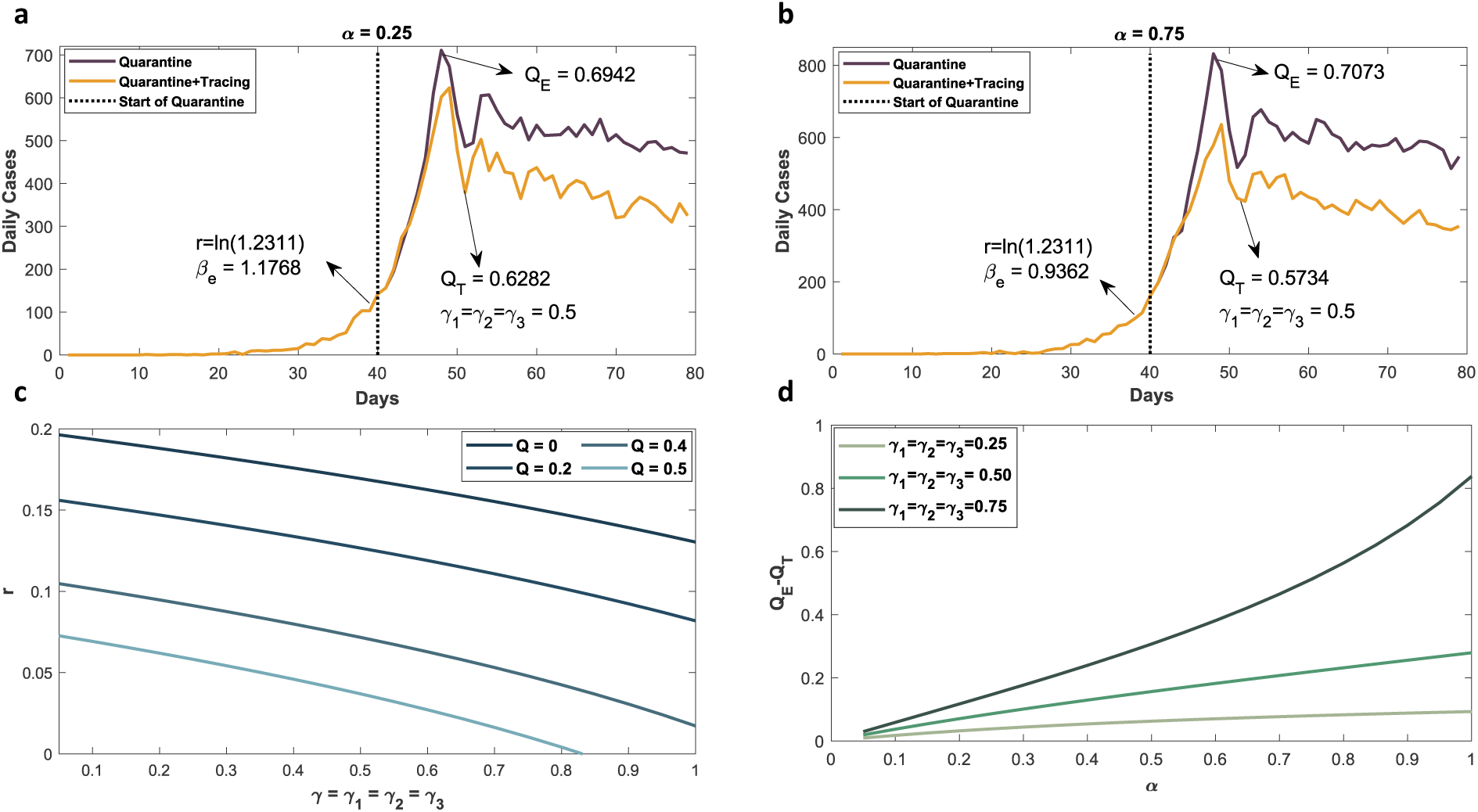
Contact tracing’s effect on decreasing the required quarantine level to stabilize the disease is highly dependent on the documentation ratio. To reach an equilibrium state, we have estimated the threshold quarantine level with (black line) and without contact tracing (golden line) from Eq. (3). The simulations starts with zero quarantine level and then proceeded with the calculated threshold quarantine level after 40 days. The initial growth rate is taken as *r* = 0.2, which is the mean value of the growth rates of countries Turkey, USA, Germany, Italy and Spain. The required *β*_*e*_ value is estimated via Eq. (2). Same simulations were made for both **a**, α = 0.25 and **b**, α = 0.75. **c**, The effect of contact tracing to the growth rate with the various values of the quarantine level. **d**, The reduction in quarantine levels calculated from equilibrium Eq. (3) with and without applying contact tracing. (see methods for parameters).

**Figure 5.**
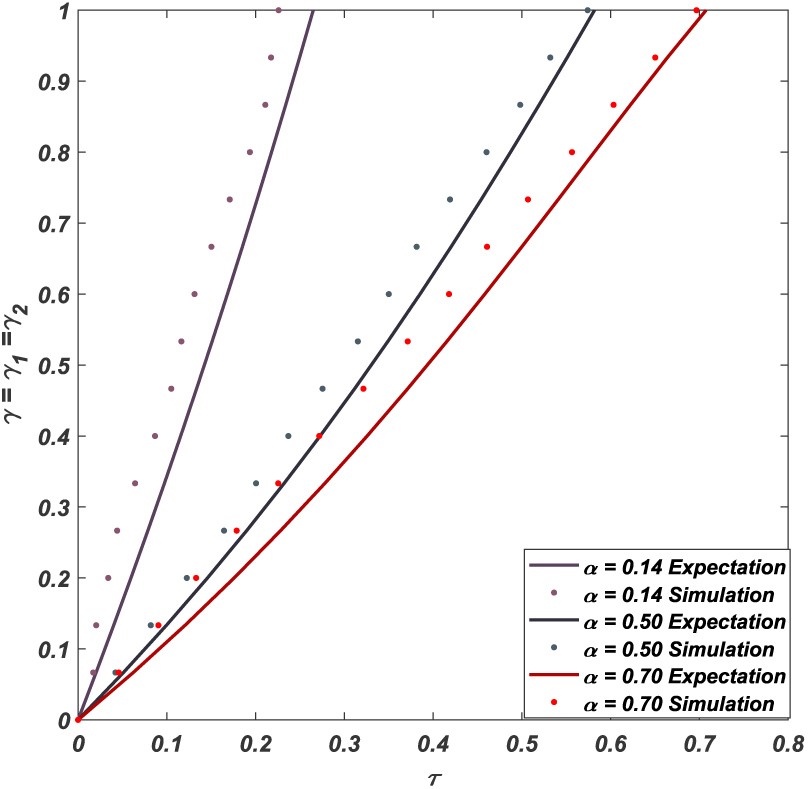
Contact tracing efficiency can be predicted by the observable known-ratio. Contact tracing efficiency was calculated for different known-ratio (*τ*). Two values of *α* = 0.14 and *α* = 0.70 were used. The known-ratio is the proportion of the newly documented patient that was being traced. For more straightforward illustration, we use the same value for all. The observable known-ratios of countries can be used as a proxy to contact tracing efficiency. The analytical solution Eq. (25) is derived from Eq. (20) assuming that pre-symptomatic state is growing/decaying exponential (see methods).

First, we have calculated the quarantine level (*Q*_*E*_ *and Q*_*T*_) through our solutions Eqs. (2) and (3). We simulate the agent-based model with the calculated parameters. The dynamics of the epidemic with the predicted (*Q*_*E*_ *and Q*_*T*_) values satisfies our expectations for both equilibrium (Fig. 4 a and 4 b) and exponential decay (Fig. S4 a and S4 b). We have seen that the outbreak reaches equilibrium after nearly 9 days of the implementation of quarantine measures. In Fig. 4 c, it is shown that how much the growth rate can be reduced by applying contact tracing under different quarantine levels for relatively low documented ratio *α* = 0.25. Since the documentation ratio is considered low, the growth rate has not significantly decreased with contact tracing. Higher documentation increases the effect of contact tracing significantly (Fig. S5). Using one of the main identities Eq. (3), we have illustrated the difference between quarantine levels with and without applying contact tracing to reach the equilibrium state (Fig. 4 d). The documentation ratio is essential to reduce the threshold quarantine level with the use of contact tracing. Extensive testing can increase the documentation ratio.

### The proportion of newly documented patients being traced from earlier patients (known-ratio) can be used to predict the contact tracing efficiency

To determine the efficiency of tracing, we have calculated the proportion of the new cases that originated from the contacts of pre-documented patients. In Fig. 6, the expected known-ratio (*τ*) calculated with Eq. (4) by using uniformly distributed parameter values of *γ*_1_ and *γ*_2_. For testing the accuracy of our estimation, the agent-based model has been simulated with the same parameters. The validation procedure is illustrated for *α* = 0.14, *α* = 0.50 and *α* = 0.70. The *α* = 0.14 is the documentation ratio found in Spain, and the *α* = 0.70 is the ratio of possibly discoverable patients found in Spain^38^. The results have been found quite predictive. Minor deviations arose because of the intrinsic stochasticity of agent-based modelling. Thus, with an analytic solution, we can calculate the efficiency of the contact-tracing from the known-ratio. A country with an observable known-ratio can predict the efficacy of their contact tracing program. The effectiveness depends mainly on the documentation ratio, which can also be observed by antibody screening.

**Figure 6.**
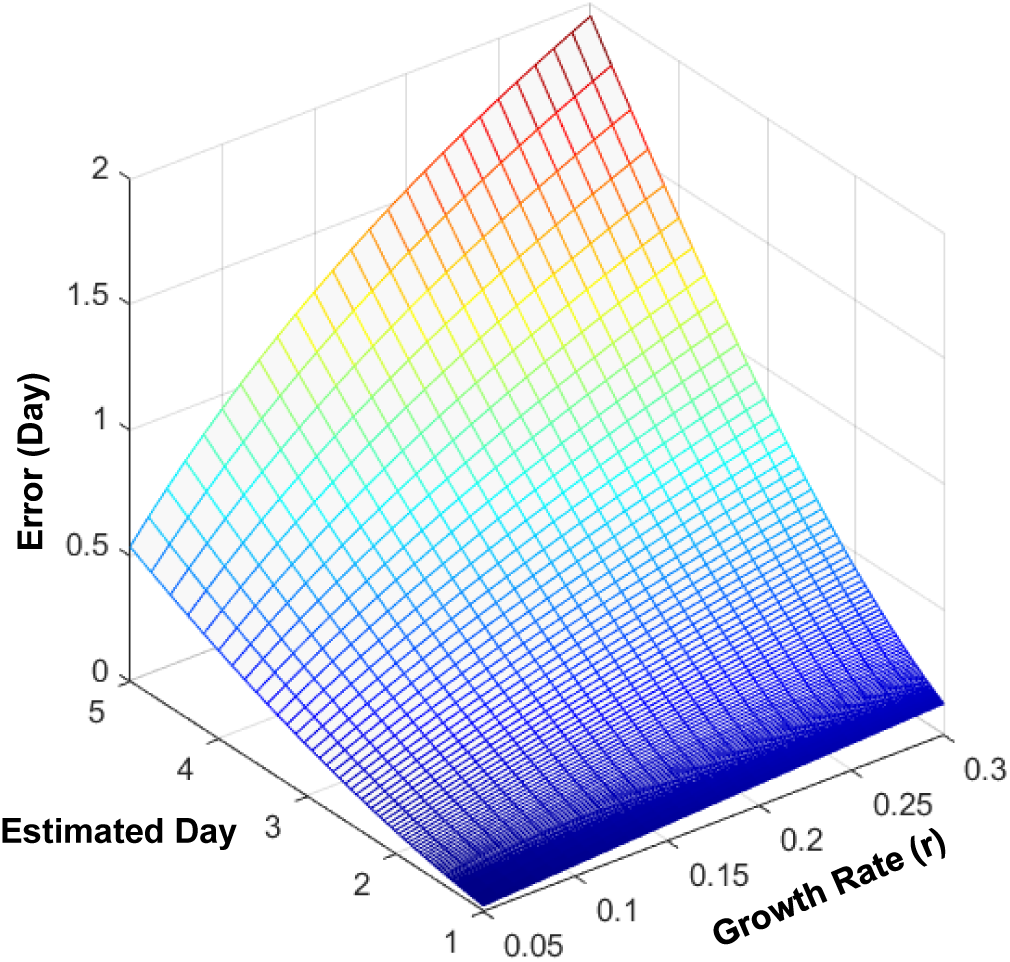
Model parameters are underestimated by uncorrected SEIR-models. Prediction of error in days when using classical SEIR-like models. The error equation is derived in the methods section as *E*(*r, day*) = |*day* − *ln*(*day* × *r* + 1)/*r*|. Higher growth rates increase underestimation.

### Uncorrected SEIR-like models underestimate the parameters of the COVID-19 dynamics

In classical SEIR-like models, the number of people who change their states is assumed as uniformly distributed. However, in the case of exponential growth or decay phases of the epidemics, the number of newly infected people is expected to be higher than the number of people who leave their stages. We have shown that the distribution of the number of people who leave their stages in compartmental models should not be uniform as adopted in SEIR-like models (for details see methods). We have calculated the error caused by accepting the uniform distribution in terms of growth rate and estimated days, as shown in Fig. 6.

As a case in point; we have used the number of daily cases in China and calculated the growth rate of the epidemics as 0.1769^32^, and obtained the latency period estimation for China as 3.69^31^ from the article that uses an SEIR-like model. The error, which indicates the miscalculation in terms of days, has been determined as nearly 1 day (Fig. 6). We have shown that the predictions of the length of the stages using the classical compartmental models are not totally correct and that our model can be used for this purpose.

## Discussion

Since the emerging of the first coronavirus cases in December 2019, several studies have attempted to understand the dynamics of the outbreak. However, the analytical relationship between the outbreak growth and other parameters, including contact tracing levels, has not been analyzed so far. In this study, we have developed a new SEIR-like model with the help of mathematical analysis of the agent-based model; and shown that it is possible to find out a novel identity which indicates the relations of the internal and external parameters of the outbreak. Our solution assumes exponential growth or decay. It is important to remember that the criteria for exponential growth or decay are low herd-immunity. At low herd-immunity, the majority of the population is susceptible. We assume susceptibles as constant, and the non-linear system of equations reduces to a linear system of differential equations. The analytical solution not only predicts the growth rate, but it can also be used to estimate parameters. Any pairwise or higher-order dependencies are easily analyzable. By contrasting SEIR-like models and agent-based model, we identified the difference between the models could be overcome by augmenting the coefficients of the model. The bridge between the two models can also be applied to different questions.

Through the linear correlation and sensitivity analyses, we have concluded that on the way of understanding the dynamics of the epidemic, correctly, it is essential to determine the roles of the infected undocumented cases as proposed by previous studies^35-37^. Sadly, we are less knowledgable about the undocumented cases. Without a doubt, this is the reason that this pandemic was never under control. The current antibody screenings show us that in Spain, 20% was documented. However, nearly 50% had or thought to have some form of symptoms. This pool might be discoverable by extensive testing or close watchfulness. If the documentation ratio was as high as 50%, with 75% effective contact tracing, the required quarantine level is only 40%. The most difference we can make is increasing the documentation ratio.

In our view, there is a big dilemma: if we assume that the documentation ratio is at max 25%, then the contact tracing should not make a meaningful difference in the growth rate; however, some countries seem to be benefiting from the contact tracing. Either the contact tracing is not a successful program, so those countries are benefiting from social distancing measures, or there is a different dynamic that we are sadly unaware.

Observing the effectiveness of the contact tracing can be difficult. We devised an equation for estimating the contact tracing effectiveness. This equation predicts the efficacy with respect to the known-ratio. The known-ratio is the ratio of the newly discovered positives from earlier traced patients to total daily positives. The known-ratio is easily observable.

One of our main findings in this study is the day estimates of SEIR-like models are not accurate because of the assumption of uniform distribution. This discrepancy can be observed in other models as well. Our analysis can also shed light into bridging SEIR-like models and agent-based models.

## Methods

### Agent-Based Model

We have developed a discrete-time stochastic agent-based model with the use of the branching process, parameterized to the COVID-19 outbreak. The branching process is established with dynamic states of *E*(*t*) (exposed), *P*(*t*) (pre-symptomatic documented), *U*(*t*) (undocumented), *D*(*t*) (symptomatic documented), *H*(*t*) (hospitalized) and *R*(*t*) (recovered) shown in Fig. 1.

The simulation process for the outbreak starts with one exposed person. Initially, an exposed agent cannot infect others during his/her latent period for *Poiss*(*d*_1_) days. After the latency period finishes, the person tends to one of the two branches; either being pre-symptomatic documented with a ratio of *α* or undocumented with a ratio of (1 − *α*). When the latency period expires, we generate a random number *K*; if *K* > *α* he/she proceeds to the undocumented stage. If the person becomes undocumented, the length of infectious period is drawn stochastically from *Poiss*(*d*_3_) distribution, he/she infects daily *Poiss*(*μβ*(1 − *Q*(*t*))) numbers of susceptible persons. When the infectious period of the undocumented stage finishes, the person goes on to the recovery stage and becomes healthy. If *K* ≤ *α* then the person proceeds to the pre-symptomatic case. The infectious period of the pre-symptomatic documented stage is *Poiss*(*d*_2_) days. Through the infectious period of the pre-symptomatic documented stage, he/she infects daily *Poiss*(*μβ*(1 − *Q*(*t*))), numbers of susceptible persons. When the infectious period of the pre-symptomatic documented stage finishes the person proceeds to the symptomatic documented stage. Person stays in the symptomatic documented stage for a random number of days, *Poiss*(*p*). Throughout the infectious period of the symptomatic documented stage, he/she infects daily *Poiss*(*β*(1 − *Q*(*t*))) numbers of susceptible persons. When the symptomatic period finishes person proceeds to the hospitalization stage where he/she will recover or die. The default parameter values for the simulations with their sources were presented in Table 1.

### Contact Tracing in the Agent-Based Model

Contact tracing starts when the person *X* proceeds to the hospitalization stage (*H*). The persons who are infected by person *X* are in the exposed stage (*E*), undocumented stage (*U*), pre-symptomatic documented stage (*P*) or symptomatic documented stage (*D*). Isolating the contacts is depending on their stages and the parameters *γ*_1_, *γ*_2_ and *γ*_3_ which are the isolation probabilities of contacts at the stage of exposed (*E*), pre-symptomatic documented (*P*) and undocumented (*U*), respectively. Contact tracing procedure in the agent-based model has been explained in the following pseudocode: begin Person *X* proceed to hospitalization stage

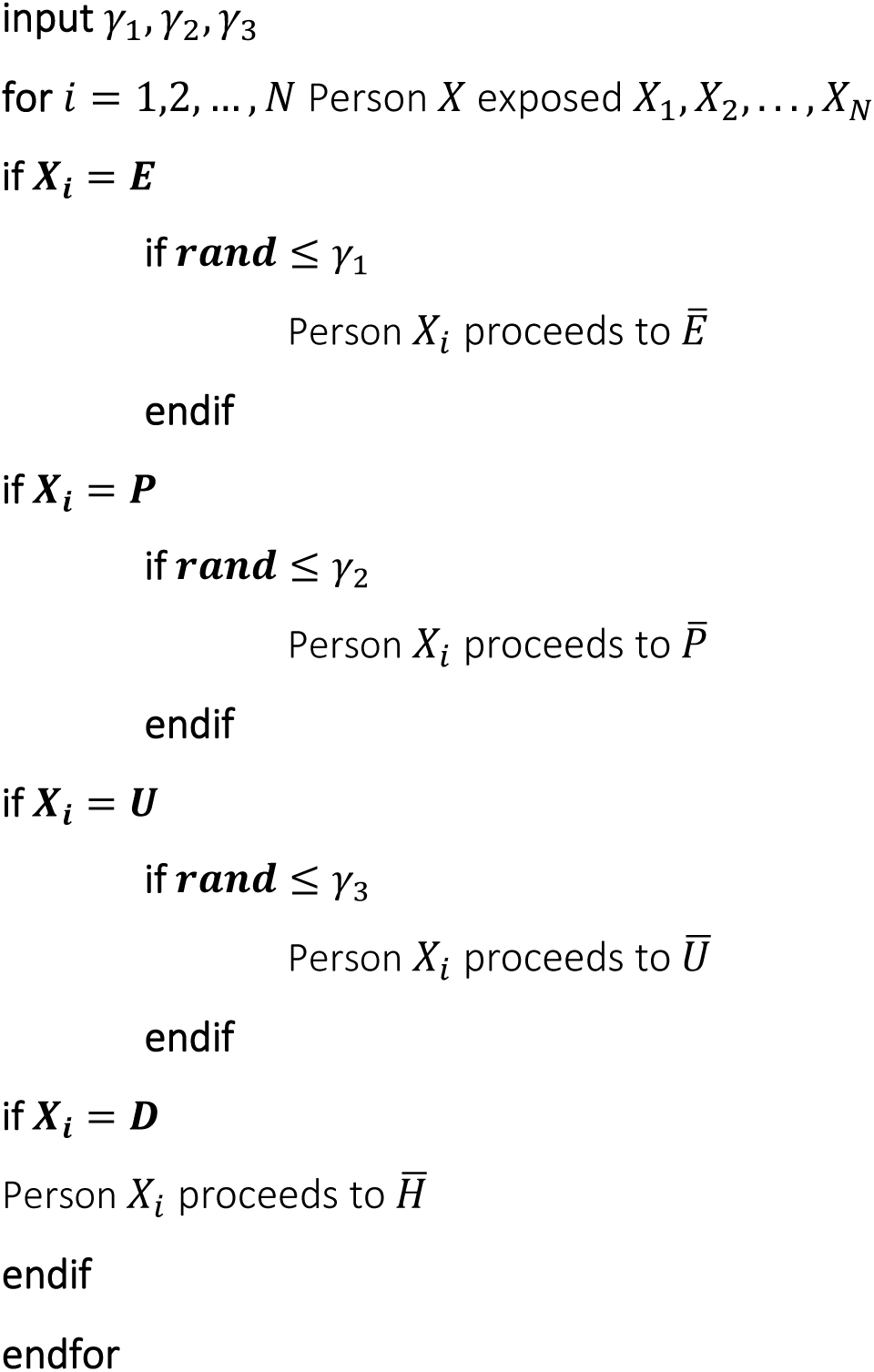

### Mathematical Analysis of the Agent-Based Model

We try to achieve a constraint that all these parameters must satisfy in the case of exponential growth or decay of the disease. Let assume that all state variables grow or decay with the rate of parameter *r*. Let us denote *E*_*x*_(*t*), *P*_*x*_(*t*), *U*_*x*_(*t*) and *D*_*x*_(*t*) as the number of people who are at these states with *x* days at any time *t*. Let us first observe that,

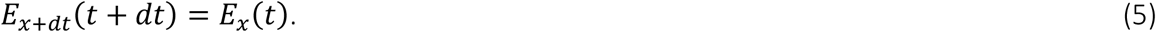

By considering exponential growth/decay of all states, we get

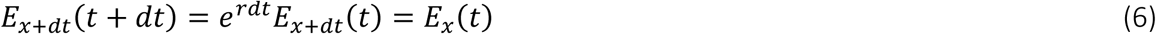

and

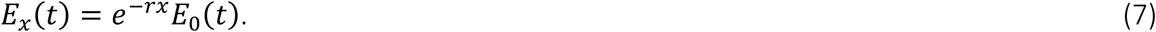

With a similar way we get

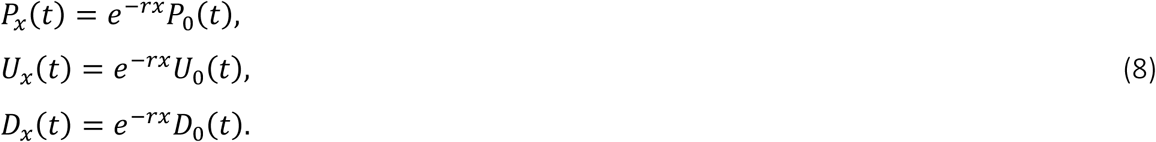

We can determine *P*_0_(*t*), *U*_0_(*t*) and *D*_0_(*t*) in terms of *E*_0_(*t*) as follows,

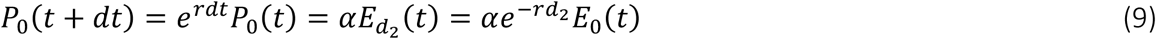

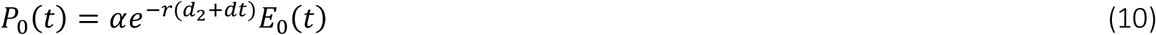

and similarly

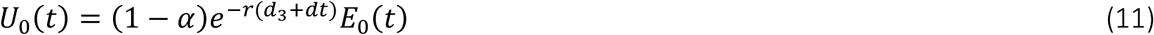

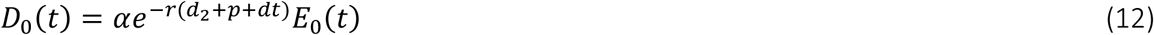

where *dt* is infinitely small time increment. New exposed individuals at *t* + *dt* can be expressed as

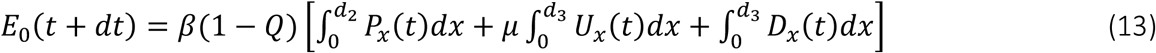

After writing the Eqs. (7)-(8) into Eq. (13) and tending *dt* to zero, we finally get the following equation,

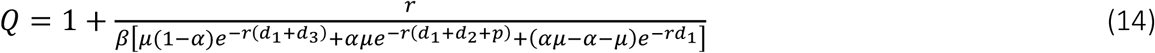

where 0 ≤ *Q* ≤ 1. The Eq. (14) implies how the internal and external parameters of the outbreak behave together. We have illustrated the relations between the growth rate and the other parameters in numerical experiments.

### The SEIR Model

The classical SEIR model with the same parameters used in the agent-based model can be stated as follows

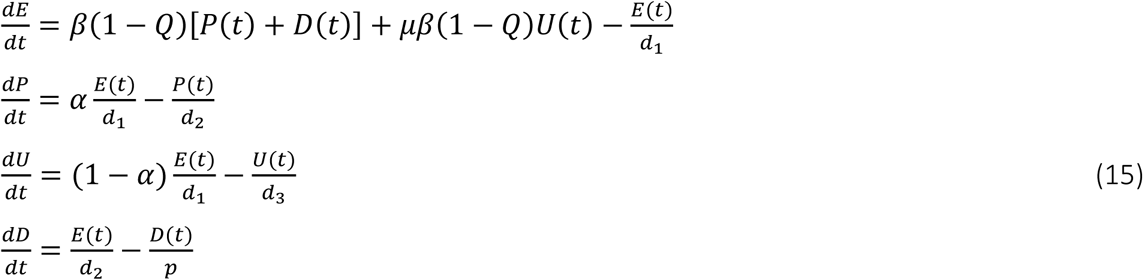

In this model, we assume that there always exist enough susceptible individuals to be infected. Additionally, since recovered and dead individuals do not affect the growth/decay dynamics, we omitted these state variables. The terms 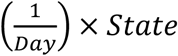 are due to the simplification of the original derivation of Kermack and Kendrick^10^. Most of the recent papers related to the modelling of COVID-19 outbreak use this simplification, and the key parameters are estimated via this assumption.

### The Corrected SEIR Model

In the equation system Eq. (15), the last terms of the differential equations indicate the uniform distribution of the state variables. For instance, the distribution of the sub-states *E*_*x*_(*t*) for 0 ≤ *x* ≤ *d*_1_ are uniform in the system Eq. (15). But as we proved in the last subsection, these statements are no longer valid when the exponential growth/decay occurs for all state variables. Thus, one needs to find out how many people should leave from his/her state at any specific time *t* for all states *E*(*t*), *P*(*t*), *U*(*t*) and *D*(*t*). With the consideration of exponential growth/decay and Eqs. (5)-(14), we get the following distributions,

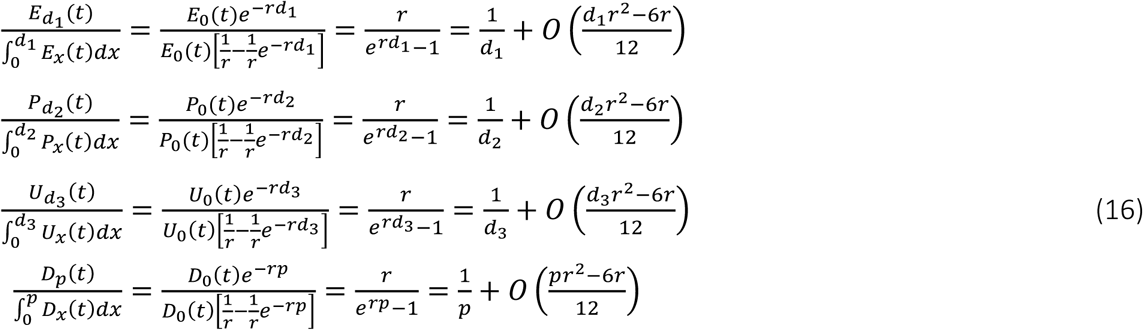

Eq. (16) show that any estimation of the exposed day, pre-symptomatic documented day, undocumented day and symptomatic documented day with the use of classical SEIR model Eq. (15) will have an approximation error. In other words, any coefficients like 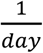 estimated with the system Eq. (15) can not exactly represent the true values of the days. Before specifying the corrected SEIR model, we can express the amount of error that will occur in the day predictions made with the classic SEIR models in the following form,

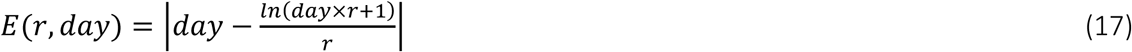

Thus, with the assumption of exponential growth/decay, the corrected SEIR model can be expressed as

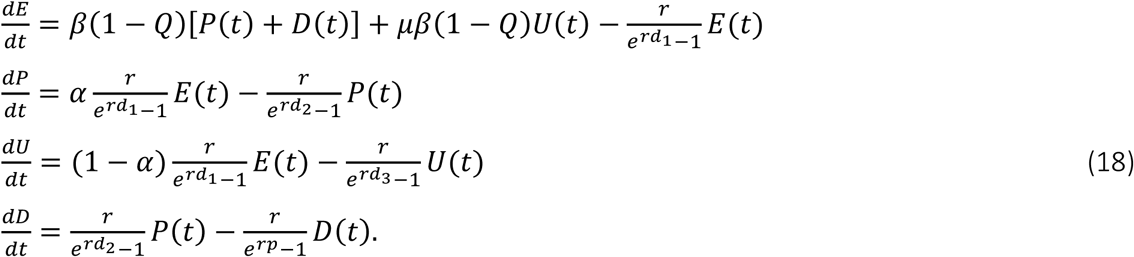

Note that, we can easily show that system Eq. (18) also satisfies Eq. (14) in exponential growth/decay. Except that the stochastic nature of the agent-based model stated in the last section, both models are equivalent if the time is continuously treated also in the agent-based model. The equilibrium equation can then be stated as

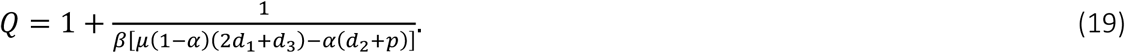

### Contact Tracing

It is important to follow the contacts of the identified cases to break the transmission chain in the COVID-19 outbreak. The growth of the outbreak can be significantly reduced by identifying and isolating people who have been contacted by people known to carry the virus. It is known from the system Eq. (17) that the number of new confirmed cases at any specific time *t* is 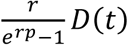. Since these group of infected cases have been transmitted the virus at the previous (*d*_2_ + *p*) days, their contacts are in the exposed stage, undocumented stage and pre-symptomatic documented stage. Here we assume that *d*_1_ > *p* and it is not possible to find out a contacted people in the symptomatic documented stage. Isolating the contacts is depending on their stage and the parameters *γ*_1_, *γ*_2_ and *γ*_3_ are used for the isolation probabilities of contacts at the stage of exposed, pre-symptomatic documented and undocumented, respectively. Thus, we get the following ODE system

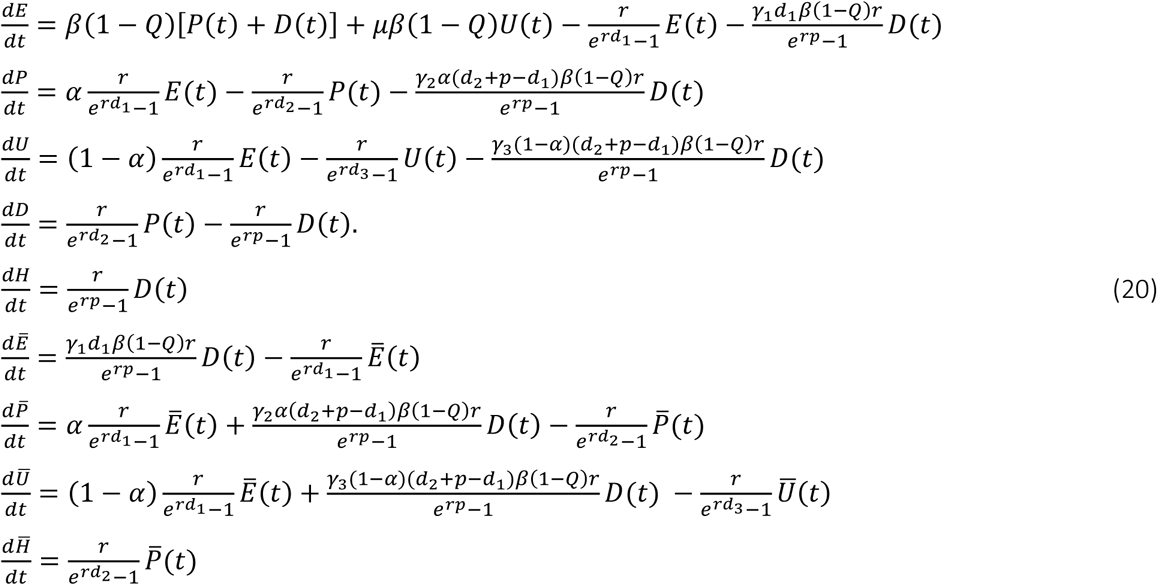

where 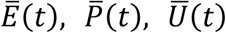 and 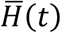 represent the isolated exposed, pre-symptomatic documented, undocumented and hospitalized number of people at any time *t*, respectively. It is important to understand how the parameters should behave for an exponential growth/decay in the presence of the contact tracing. Applying a similar procedure by considering *P*(*t*) = *P*_0_*e*^*rt*^, the desired growth/decay functions of the other states in large time can be obtained as

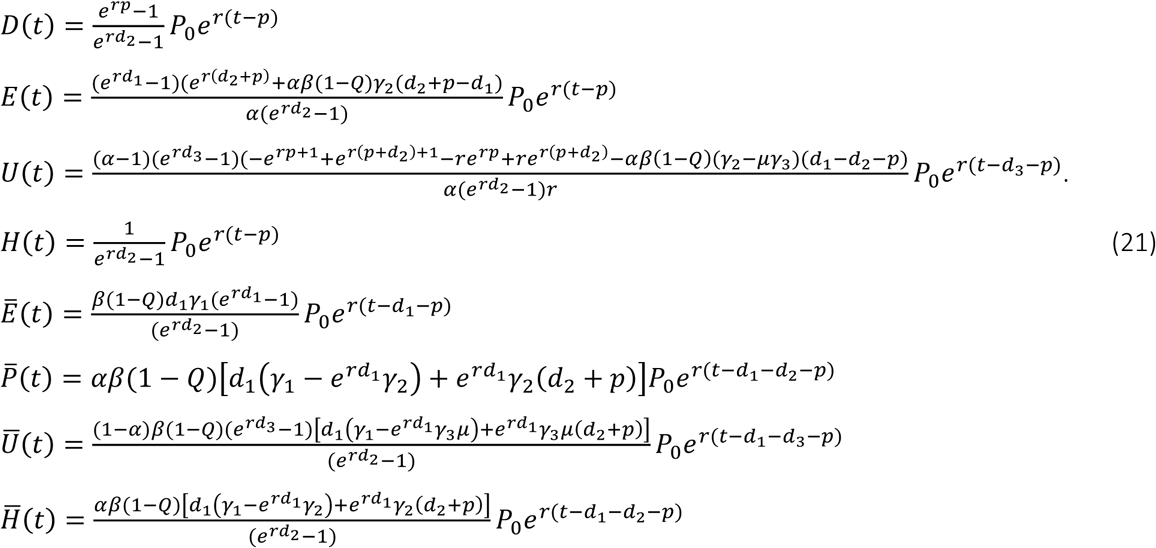

Finally, the following constraint can be achieved for the system Eqs. (20)

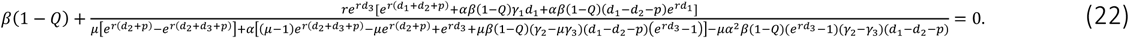

The equilibrium state equation can be derived from Eq. (22) as follows,

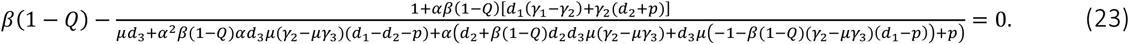

The efficiency of the applied contact tracing can be measured by the evaluation of the following known-ratio:

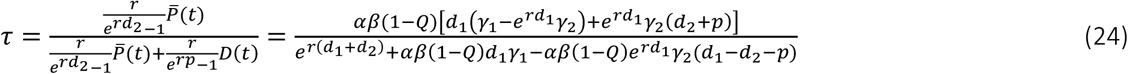

i.e. the rate of isolated hospitalized cases in the total hospitalized number of people at any time *t*. With the consideration of equal tracing probabilities *γ* = *γ*_1_ = *γ*_2_ = *γ*_3_, the contact tracing probability *γ* can be written in terms of the known-ratio and other parameters as follows

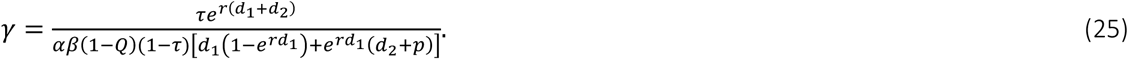

### Parameters

The model parameters and default values have been demonstrated in Table 1. Depending on the given figures we can summarize the parameter values as follows;

*Figure 2. d*_1_ = 3.69, *d*_2_ = 3.47, *d*_3_ = 3.47, *p* = 1.92, *α* = 0.14, *β* = 1.12, *μ* = 0.55, *Q* = 0, *γ*_1_ = *γ*_2_ = *γ*_3_ = 0. Together with these default parameter values, all parameters individually changed in subfigures as illustrated in *x* −axis.

*Figure 3 a*. The parameter values are randomly selected from the intervals 0.5 ≤ *d*_1_ ≤ 5, 0.5 ≤ *d*_2_ ≤ 5, 0.5 ≤ *d*_3_ ≤ 5, 0 ≤ *p* ≤ 5, 0 ≤ *α* ≤ 1, 0 ≤ *β* ≤ 5, 0 ≤ *μ* ≤ 1, 0 ≤ *γ*_1_ ≤ 1, 0 ≤ *γ*2 ≤ 1, 0 ≤ *γ*3 ≤ 1. 10,000 different set of parameters taken and the corresponding growth rates are evaluated from novel Eq. (2).

*Figure 3 b. d*_1_ = 3.69, *d*_2_ = 3.47, *d*_3_ = 3.47, *p* = 1.92, *α* = 0.14, *β* = 1.12, *μ* = 0.55, *Q* = 0, *γ*_1_ = *γ*_2_ = *γ*_3_ = 0. Together with these default parameter values, the growth rate is evaluated from novel Eq. (2) as *r* = 0.1767.

*Figure 3 c. d*_1_ = 3.69, *d*_2_ = 3.47, *d*_3_ = 3.47, *p* = 1.92, *β* = 1.12, *μ* = 0.55, *Q* = 0, *γ*_1_ = *γ*_2_ = *γ*_3_ = 0. Together with these default parameter values, the growth rates are evaluated from novel Eq. (2) with respect to the changing values of *α*.

*Figure 3 d. d*_1_ = 3.69, *d*_2_ = 3.47, *d*_3_ = 3.47, *p* = 1.92, *β* = 1.12, *μ* = 0.55, *Q* = 0, *γ*_1_ = *γ*_2_ = *γ*_3_ = 1. Together with these default parameter values, the growth rates are evaluated from novel Eq. (2) with respect to the changing values of *α*.

*Figure 4 a*.

Simulations without applying contact tracing. Before quarantine application: *d*_1_ = 3.69, *d*_2_ = 3.47, *d*_3_ = 3.47, *p* = 1.92, *α* = 0.25, *β* = 1.17, *μ* = 0.55, *Q* = 0, *r* = 0.2, *γ*_1_ = *γ*_2_ = *γ*_3_ = 0. After quarantine application: *d*_1_ = 3.69, *d*_2_ = 3.47, *d*_3_ = 3.47, *p* = 1.92, *α* = 0.14, *β* = 1.17, *μ* = 0.55, *Q* = 0.6942, *r* = 0, *γ*_1_ = *γ*_2_ = *γ*_3_ = 0.

Simulations with applying contact tracing. Before quarantine application: *d*_1_ = 3.69, *d*_2_ = 3.47, *d*_3_ = 3.47, *p* = 1.92, *α* = 0.25, *β* = 1.17, *μ* = 0.55, *Q* = 0, *r* = 0.2, *γ*_1_ = *γ*_2_ = *γ*_3_ = 0. After quarantine application: *d*_1_ = 3.69, *d*_2_ = 3.47, *d*_3_ = 3.47, *p* = 1.92, *α* = 0.14, *β* = 1.17, *μ* = 0.55, *Q* = 0.6282, *r* = 0, *γ*_1_ = *γ*_2_ = *γ*_3_ = 0.5.

*Figure 4 b*.

Simulations without applying contact tracing. Before quarantine application: *d*_1_ = 3.69, *d*_2_ = 3.47, *d*_3_ = 3.47, *p* = 1.92, *α* = 0.75, *β* = 0.93, *μ* = 0.55, *Q* = 0, *r* = 0.2, *γ*_1_ = *γ*_2_ = *γ*_3_ = 0. After quarantine application: *d*_1_ = 3.69, *d*_2_ = 3.47, *d*_3_ = 3.47, *p* = 1.92, *α* = 0.75, *β* = 0.93, *μ* = 0.55, *Q* = 0.7073, *r* = 0, *γ*_1_ = *γ*_2_ = *γ*_3_ = 0.

Simulations with applying contact tracing. Before quarantine application: *d*_1_ = 3.69, *d*_2_ = 3.47, *d*_3_ = 3.47, *p* = 1.92, *α* = 0.75, *β* = 0.93, *μ* = 0.55, *Q* = 0, *r* = 0.2, *γ*_1_ = *γ*_2_ = *γ*_3_ = 0. After quarantine application: *d*_1_ = 3.69, *d*_2_ = 3.47, *d*_3_ = 3.47, *p* = 1.92, *α* = 0.75, *β* = 0.93, *μ* = 0.55, *Q* = 0.6282, *r* = 0, *γ*_1_ = *γ*_2_ = *γ*_3_ = 0.5.

*Figure 4 c. d*_1_ = 3.69, *d*_2_ = 3.47, *d*_3_ = 3.47, *p* = 1.92, *α* = 0.25, *β* = 1.12, *μ* = 0.55, *Q* = 0. *Figure 4 d. d*_1_ = 3.69, *d*_2_ = 3.47, *d*_3_ = 3.47, *p* = 1.92, *α* = 0.25, *β* = 1.12, *μ* = 0.55, *Q* = 0, *r* = 0.2.

*Figure 5. d*_1_ = 3.69, *d*_2_ = 3.47, *d*_3_ = 3.47, *p* = 1.92, *α* = 0.14/0.70, *β* = 1.12, *μ* = 0.55, *Q* = 0, *γ*_1_ = *γ*_2_ = *γ*_3_ = 0. Together with these default parameter values, the growth rate is evaluated from novel Eq. (2) with respect to changing values of contact tracing probabilities.

*Figure S1. d*_1_ = 3.69, *d*_2_ = 3.47, *d*_3_ = 3.47, *p* = 1.92, *α* = 0.14, *r* = 0.1767, *μ* = 0.55, *Q* = 0, *γ*_1_ = *γ*_2_ = *γ*_3_ = 0. Together with these default parameter values, all parameters individually changed in subfigures as illustrated in *x* −axis. *β* values are obtained from novel Eq. (2).

*Figure S2. d*1 = 3.69, *d*2 = 3.47, *d*3 = 3.47, *p* = 1.92, *α* = 0.25, *β* = 1.12, *μ* = 0.55, *Q* = 0, *γ*_1_ = *γ*_2_ = *γ*_3_ = 0.75. Together with these default parameter values, all parameters individually changed in subfigures as illustrated in *x* −axis. *r* values are obtained from novel Eq. (2) and known-ratio (*τ*) values are obtained from Eq. (4).

*Figure S3. d*_1_ = 3.69, *d*_2_ = 3.47, *d*_3_ = 3.47, *p* = 1.92, *r* = 0.2, *μ* = 0.55. Together with these default parameter values, the documented ratio is taken uniformly as illustrated in *x* −axis. *β* values are obtained from novel Eq. (2) and *Q*_*T*_ values are obtained from Eq. (3).

*Figure S4 a*.

Simulations without applying contact tracing. Before quarantine application: *d*_1_ = 3.69, *d*_2_ = 3.47, *d*_3_ = 3.47, *p* = 1.92, *α* = 0.25, *β* = 1.17, *μ* = 0.55, *Q* = 0, *r* = 0.2, *γ*_1_ = *γ*_2_ = *γ*_3_ = 0. After quarantine application: *d*_1_ = 3.69, *d*_2_ = 3.47, *d*_3_ = 3.47, *p* = 1.92, *α* = 0.14, *β* = 1.17, *μ* = 0.55, *Q* = 0.9139, *r* = −0.2, *γ*_1_ = *γ*_2_ = *γ*_3_ = 0.

Simulations with applying contact tracing. Before quarantine application: *d*_1_ = 3.69, *d*_2_ = 3.47, *d*_3_ = 3.47, *p* = 1.92, *α* = 0.25, *β* = 1.17, *μ* = 0.55, *Q* = 0, *r* = 0.2, *γ*_1_ = *γ*_2_ = *γ*_3_ = 0. After quarantine application: *d*_1_ = 3.69, *d*_2_ = 3.47, *d*_3_ = 3.47, *p* = 1.92, *α* = 0.14, *β* = 1.17, *μ* = 0.55, *Q* = 0.8668, *r* = −0.2, *γ*_1_ = *γ*_2_ = *γ*_3_ = 0.5.

*Figure S4 b*.

Simulations without applying contact tracing. Before quarantine application: *d*_1_ = 3.69, *d*_2_ = 3.47, *d*_3_ = 3.47, *p* = 1.92, *α* = 0.75, *β* = 0.93, *μ* = 0.55, *Q* = 0, *r* = 0.2, *γ*_1_ = *γ*_2_ = *γ*_3_ = 0. After quarantine application: *d*_1_ = 3.69, *d*_2_ = 3.47, *d*_3_ = 3.47, *p* = 1.92, *α* = 0.75, *β* = 0.93, *μ* = 0.55, *Q* = 0.9221, *r* = −0.2, *γ*_1_ = *γ*_2_ = *γ*_3_ = 0.

Simulations with applying contact tracing. Before quarantine application: *d*_1_ = 3.69, *d*_2_ = 3.47, *d*_3_ = 3.47, *p* = 1.92, *α* = 0.75, *β* = 0.93, *μ* = 0.55, *Q* = 0, *r* = 0.2, *γ*_1_ = *γ*_2_ = *γ*_3_ = 0. After quarantine application: *d*_1_ = 3.69, *d*_2_ = 3.47, *d*_3_ = 3.47, *p* = 1.92, *α* = 0.75, *β* = 0.93, *μ* = 0.55, *Q* = 0.8379, *r* = −0.2, *γ*_1_ = *γ*_2_ = *γ*_3_ = 0.5.

*Figure S5. d*_1_ = 3.69, *d*_2_ = 3.47, *d*_3_ = 3.47, *p* = 1.92, *α* = 0.70, *μ* = 0.55, *β* = 1.12. Together with these default parameter values, contact tracing probabilities are taken uniformly as illustrated in *x* −axis. *r* values are obtained from novel Eq. (2).

## Data Availability

Codes are available by request

## Author Information

### Contributions

H.T. methodology, simulations, formal analysis, conceptualization,

F.Z.S writing—original draft;

B.N.D. visualization, writing—original draft;

R.N. methodology;

M.S. supervision, editing.

S.K. supervision, methodology, formal analysis, conceptualization, writing-reviewing and editing;

