## Supplemental Figures for "Analytical solution of equivalent SEIR and agent-based model of COVID-19; showing the bounds of contact tracing"

With the help of our novel Eq. (2), the transmission rate of documented cases $(\beta)$, which is one of the key parameters of the COVID-19 dynamics, has been calculated with respect to changing values of the other model parameters (Fig. S1). The expected known-rate $(\tau)$, which is the ratio of the newly discovered positives from earlier traced patients to total daily positives, can be evaluated from Eq. (4) as a multidimensional function of model parameters. Once the default parameters are accepted (see Fig. 1), we have shown how the known-rate $(\tau)$ changes according to each model parameter in Fig. S2.

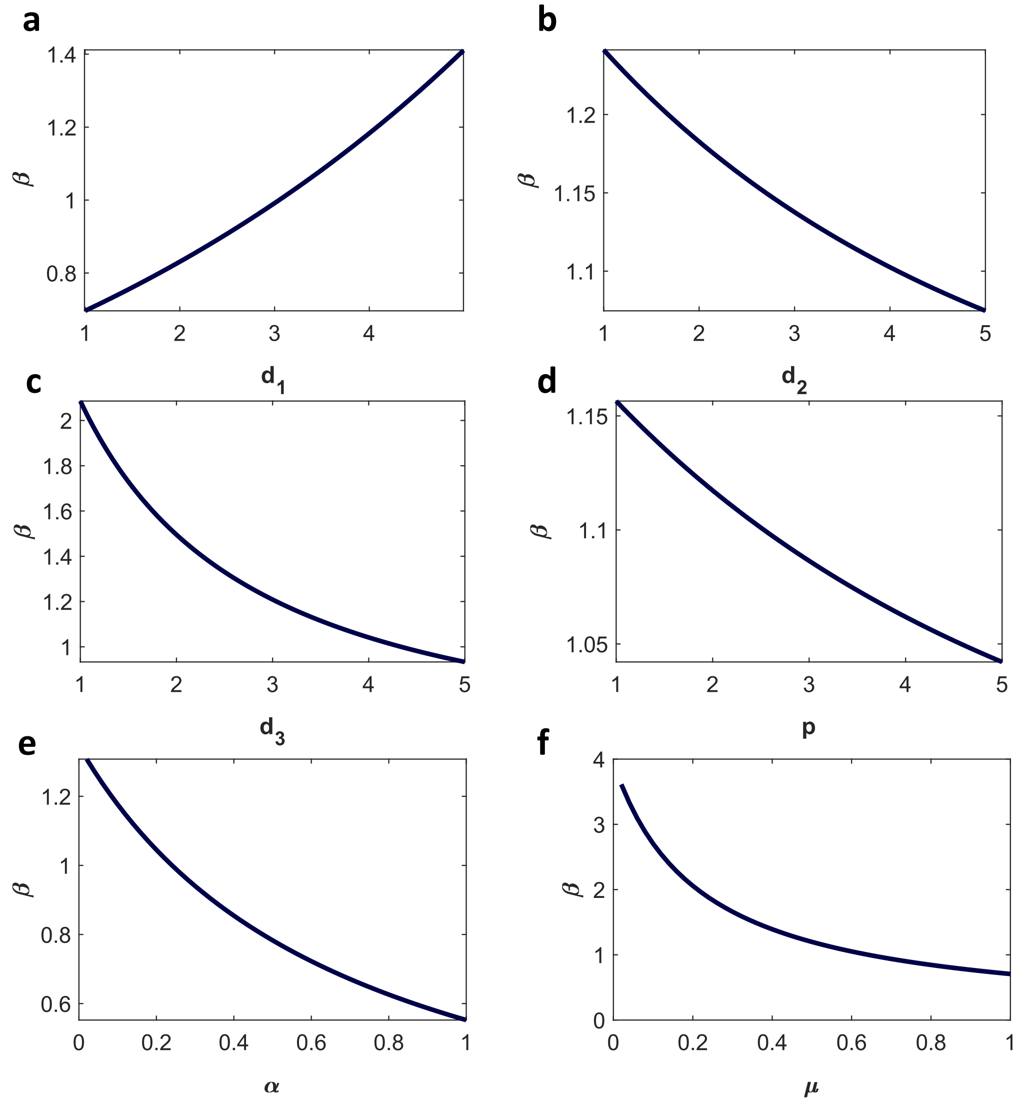


**Figure S1.** The change of the transmission rate of documented cases $(\beta)$ with respect to **a**, the mean number of days in exposed stage $d_1$ **b**, the mean number of days in the undocumented stage $d_2$ **c**, the mean number of days in the pre-symptomatic documented stage $d_3$ **d**, the mean number of days in the symptomatic documented stage $p$ **e**, the fraction of documented cases $\alpha$ **f**, transmission reduction factor for undocumented cases $\mu$.

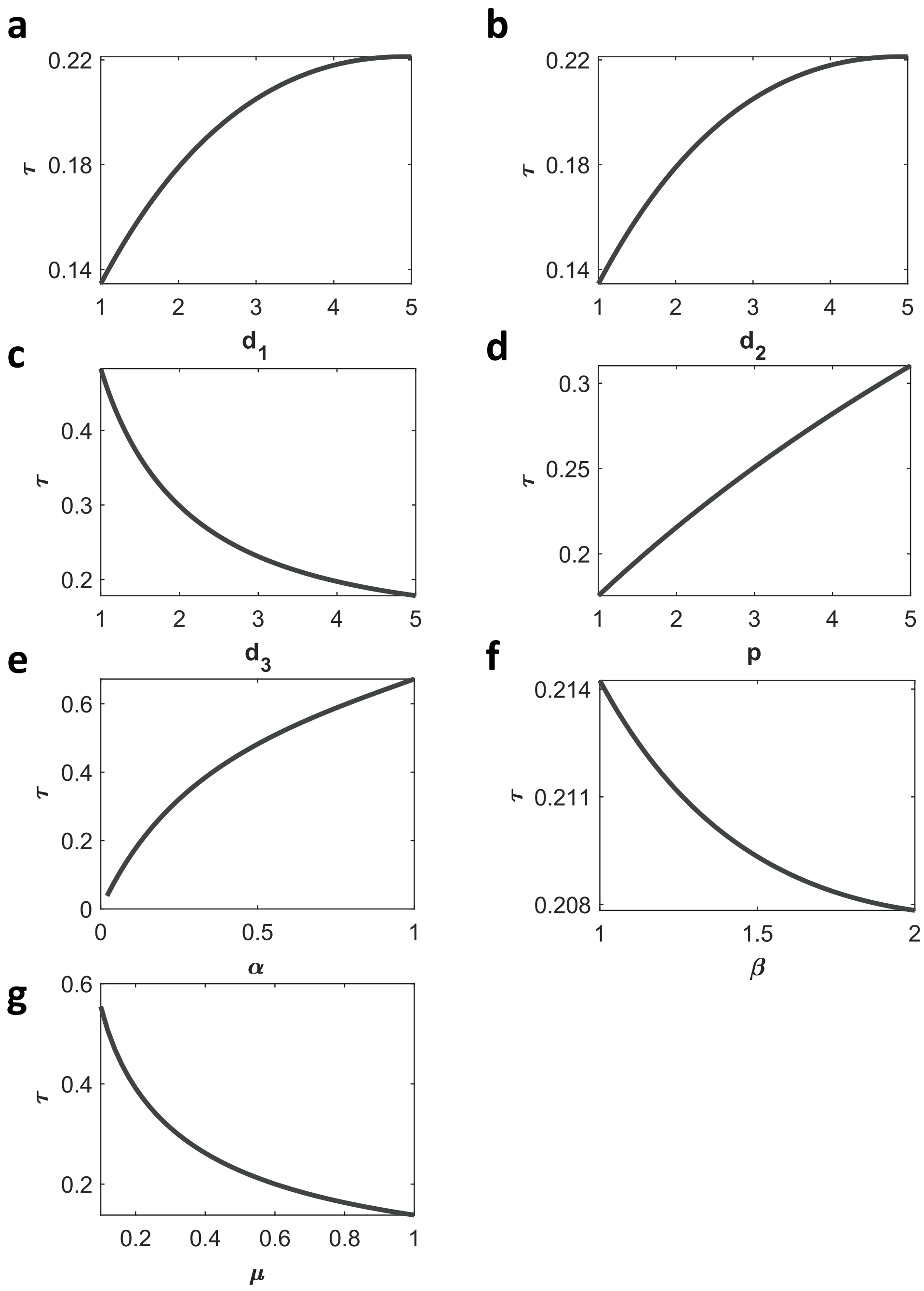


Figure S2. In the presence of 75% contact tracing rate ($\gamma_1 = \gamma_2 = \gamma_3 = 0.75$), the change of the known rate ($\tau$) with respect to **a**, the mean number of days in exposed stage $d_1$ **b**, the mean number of days in the pre-symptomatic documented stage $d_2$ **c**, the mean number of days in the undocumented stage $d_3$ **d**, the mean number of days in the symptomatic documented stage $p$ **e**, the fraction of documented cases $\alpha$ **f**, infectiousness rate of the symptomatic documented cases $\beta$ **g**, transmission reduction factor for undocumented cases $\mu$.

The quarantine rates required to reach equilibrium in the presence of different tracing rates and documented rates have been illustrated in Fig. S3 with the use of novel Eq. (3).

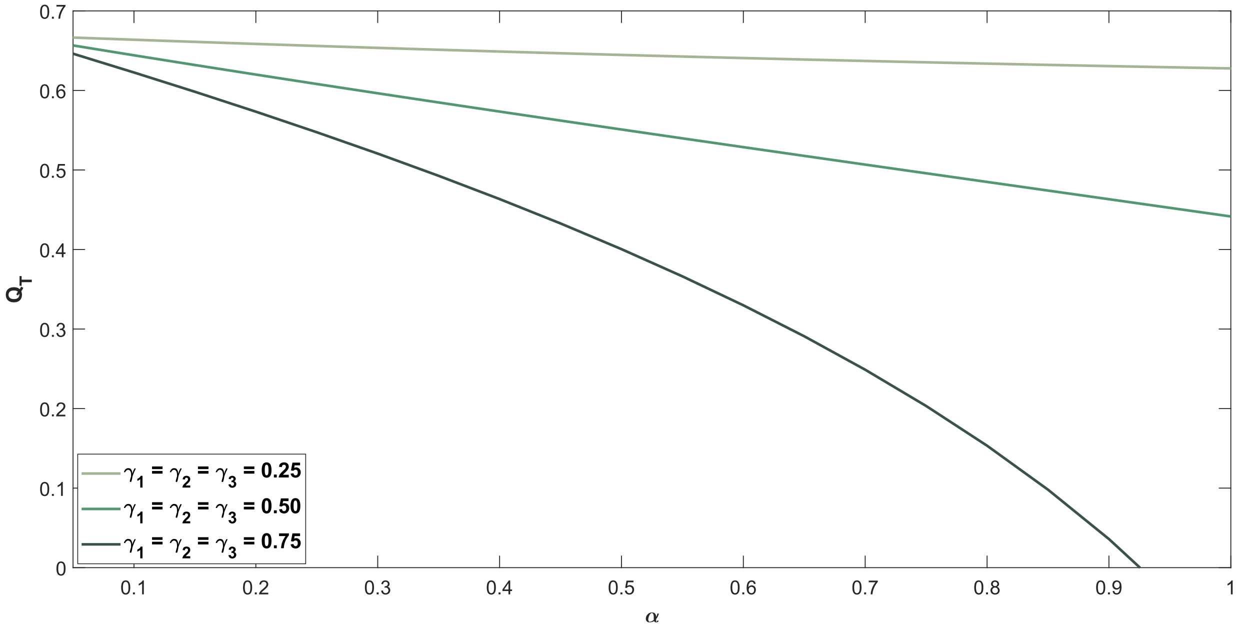


**Figure S3.** The threshold quarantine levels to reach equilibrium. The curves are obtained with respect to various values of the documentation ratio $\alpha$ and contact tracing rates.

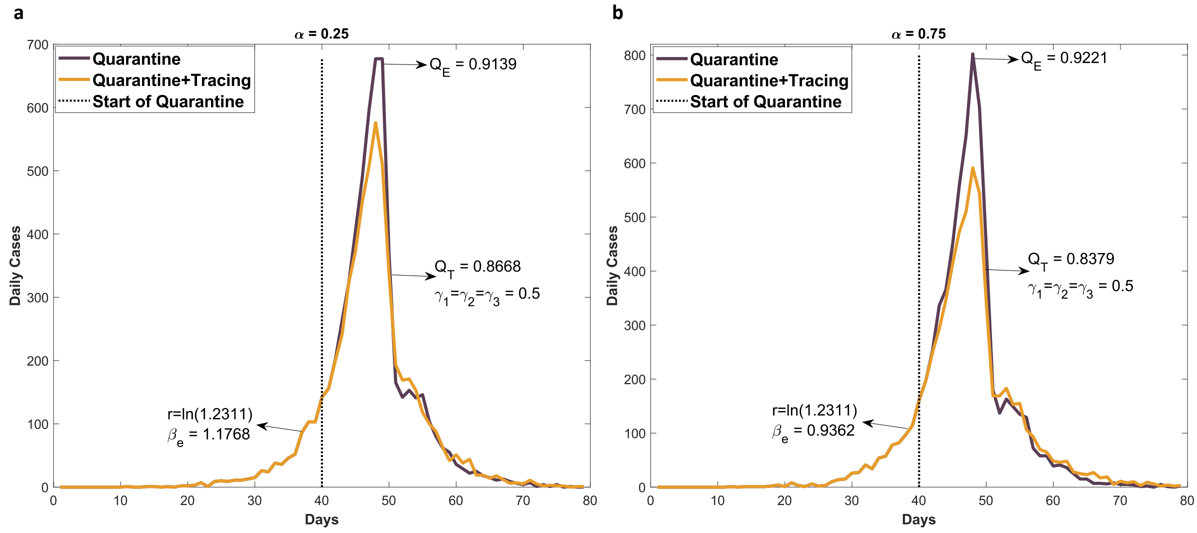


**Figure S4.** Estimations of the quarantine rates for specified exponential decay and efficiency of contact tracing. The growth rate is taken as $r = 0.2$, which is the mean value of the growth rates of countries Turkey, USA, Germany, Italy and Spain. The required $\beta_e$ value is estimated for both **a,** $\alpha = 0.25$ and **b**, $\alpha = 0.75$ via Eq. (2). The agent-based simulation is performed with these parameters for up to 40 days. To exponentially decay with the decay rate r=-0.2, we have estimated the threshold quarantine rates with and without applying contact tracing from Eq. (2) for both $\alpha = 0.25$ and $\alpha = 0.75$, respectively. The simulations have proceeded with these threshold quarantine rates after 40 days.

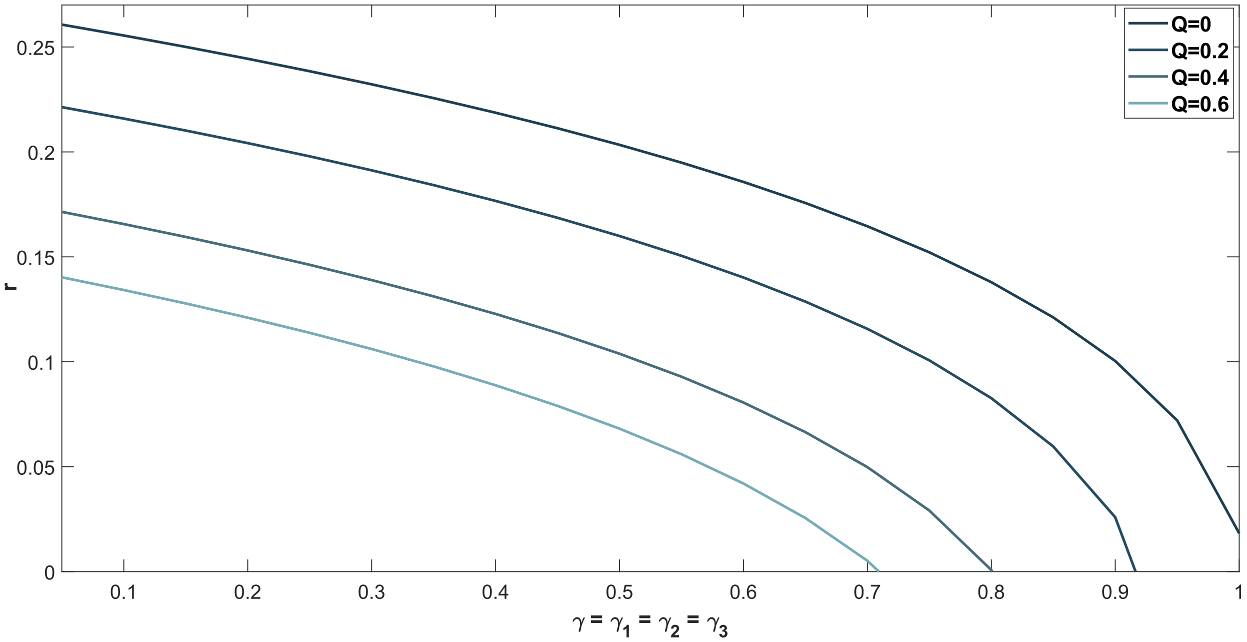


**Figure S5.** The effect of contact tracing to the growth rate in the presence of the various values of the quarantine rates.
